# Stage-Dependent Differences in Quality of Life Among Breast Cancer Patients Prior to Systemic Therapy: A Cross-sectional Study

**DOI:** 10.64898/2025.12.01.25341389

**Authors:** Henry Sutanto, Merlyna Savitri, Een Hendarsih, Ami Ashariati

## Abstract

**Background:** Disease stage is a major determinant of health-related quality of life (HRQoL) in breast cancer, yet stage-stratified HRQoL data from low-and middle-income settings remain limited. Understanding baseline differences across stages is essential for optimizing supportive care before systemic therapy.

**Methods:** In this cross-sectional study conducted at two secondary referral centers in East Java, Indonesia, from January to October 2025, we consecutively enrolled women with histologically confirmed breast cancer prior to initiation of systemic therapy. Demographic and tumor characteristics were recorded, including histopathology, grade, and immunohistochemistry. HRQoL was assessed using validated versions of the EORTC QLQ-C30 and QLQ-BR23. Patients were categorized into early (n=14), locally advanced (n=62), and metastatic disease (n=30). Group differences were analyzed using the Kruskal–Wallis test.

**Results:** Participants were exclusively female, with slightly younger age in advanced and metastatic groups. Invasive ductal carcinoma predominated across stages (80–100%). Higher-grade tumors were most frequent in locally advanced disease, while Luminal B and HER2-enriched subtypes were more common in advanced stages. HRQoL demonstrated clear stage-dependent decline. Global health status was highest in early disease (median 83.3) and lowest in metastatic disease (75.0; *p* = 0.004). Social functioning differed significantly across stages (*p* = 0.022). Symptom burden—including pain (*p* = 0.041) and systemic side effects (*p* = 0.025)—was greatest in metastatic disease. Emotional, cognitive, and body-image domains were similar between groups.

**Conclusion:** Breast cancer stage is strongly associated with baseline HRQoL impairment, particularly in global health, social functioning, pain, and treatment-related symptoms. These findings highlight the need for earlier diagnosis and stage-specific supportive care strategies in resource-constrained settings.

## Introduction

Breast cancer remains the most commonly diagnosed malignancy among women worldwide and continues to impose substantial morbidity, mortality, and psychosocial burden [1]. Although therapeutic advancements have markedly improved survival—particularly in high-income countries—disease stage at diagnosis remains a dominant determinant of clinical outcomes and patient experience. In many low-and middle-income regions, including large parts of Southeast Asia, women frequently present with locally advanced or metastatic disease due to delayed detection, restricted access to screening, sociocultural barriers, and variable health literacy [2]. These stage-related disparities not only influence survival probabilities but also profoundly shape symptom burden, functional limitations, and health-related quality of life (HRQoL) even before systemic therapy begins. HRQoL has emerged as an indispensable clinical endpoint in breast cancer care, complementing traditional biomarkers and survival metrics [3]. The European Organisation for Research and Treatment of Cancer (EORTC) QLQ-C30 and QLQ-BR23 questionnaires—robust, validated tools employed globally—allow clinicians and researchers to quantify multidimensional aspects of patient well-being, including physical, emotional, cognitive, and social functioning, as well as specific symptoms such as pain, fatigue, and systemic effects [4]. However, despite the widespread use of these instruments internationally, there is limited stage-stratified HRQoL evidence from real-world populations in low-resource settings, where patients often present with more aggressive tumor biology and heavier symptom burdens. As such, global literature may underestimate the HRQoL challenges faced by patients in regions where late presentation is the norm rather than the exception.

Beyond HRQoL itself, an important clinical gap relates to the integration of baseline patient-reported outcomes (PROs) into treatment planning. Pre-treatment QoL provides a snapshot of patient vulnerability—particularly in advanced disease—informing decisions about the intensity of systemic therapy, the need for early supportive interventions, and realistic prognostication [5]. Advanced-stage patients typically endure higher symptom loads due to tumor burden, systemic inflammation, and psychosocial distress, while early-stage patients may experience anxiety related to diagnosis but fewer physical limitations [6,7]. Yet, the magnitude and character of these stage-dependent differences remain underexplored in many parts of the world. Without such data, health systems cannot develop comprehensive supportive-care frameworks that reflect local disease epidemiology and patient needs. This research was designed with the overarching goal of characterizing the stage-specific differences in baseline HRQoL among women with breast cancer prior to initiating systemic therapy. Specifically, the study aimed to describe the demographic, clinical, histopathologic, and molecular characteristics of patients stratified by early, locally advanced, and metastatic breast cancer; assess baseline HRQoL across multiple functional and symptom domains using validated EORTC QLQ-C30 and QLQ-BR23 instruments; compare HRQoL outcomes across disease stages to determine which aspects of patient well-being are most affected by advancing tumor burden; identify symptom clusters and functional deficits that may necessitate stage-specific supportive interventions prior to pharmacotherapy. Through these aims, the study provides globally relevant evidence on how breast cancer stage shapes pre-treatment patient experience, with direct implications for clinical decision-making, supportive care, and health-system planning.

## Materials and Methods

### Study design

This was a cross-sectional study conducted at two secondary referral centers in East Java, Indonesia, from January to October 2025, both of which provide comprehensive diagnostic and therapeutic services for breast cancer. The study was designed to evaluate baseline HRQoL across different clinical stages of breast cancer prior to the initiation of systemic therapy. Data collection occurred during each patient’s pre-treatment evaluation, ensuring that HRQoL assessments reflected the impact of disease itself rather than treatment-induced toxicity. Ethical approvals were obtained from the institutional review boards of both participating hospitals (197/KEP/2024 from Universitas Airlangga Hospital and 445/02/KOM.ETIK/2025 from Haji General Hospital), and all participants provided written informed consent.

### Study population and sample selection

The study consecutively enrolled adult women with histologically confirmed breast cancer who were scheduled to begin systemic pharmacotherapy, including chemotherapy, endocrine therapy, or targeted therapy. Patients were recruited consecutively during outpatient oncology evaluations to reflect real-world clinical presentation. Based on prior regional epidemiological estimates and HRQoL variance data [4,8], a minimum sample size of 90 participants was calculated to ensure adequate statistical power for stage-stratified comparisons. Ultimately, 106 patients were included and categorized into early-stage, locally advanced, and metastatic groups according to TNM-based clinical staging.

### Inclusion and exclusion criteria

Eligible participants were women aged 18 years or older with histopathologically confirmed breast cancer (**Figure 1**). All participants were required to be able to understand and complete the HRQoL questionnaires and have a planned initiation of systemic therapy following baseline assessment. Patients were excluded if they had severe cognitive impairment or psychiatric conditions that could interfere with questionnaire reliability, a concurrent active malignancy, recent systemic treatment within the last 3 months that could affect baseline HRQoL values, or if they were unwilling or unable to provide informed consent. These criteria ensured that the baseline assessments captured the true impact of disease burden rather than residual treatment effects or unrelated clinical factors.

**Figure 1.**
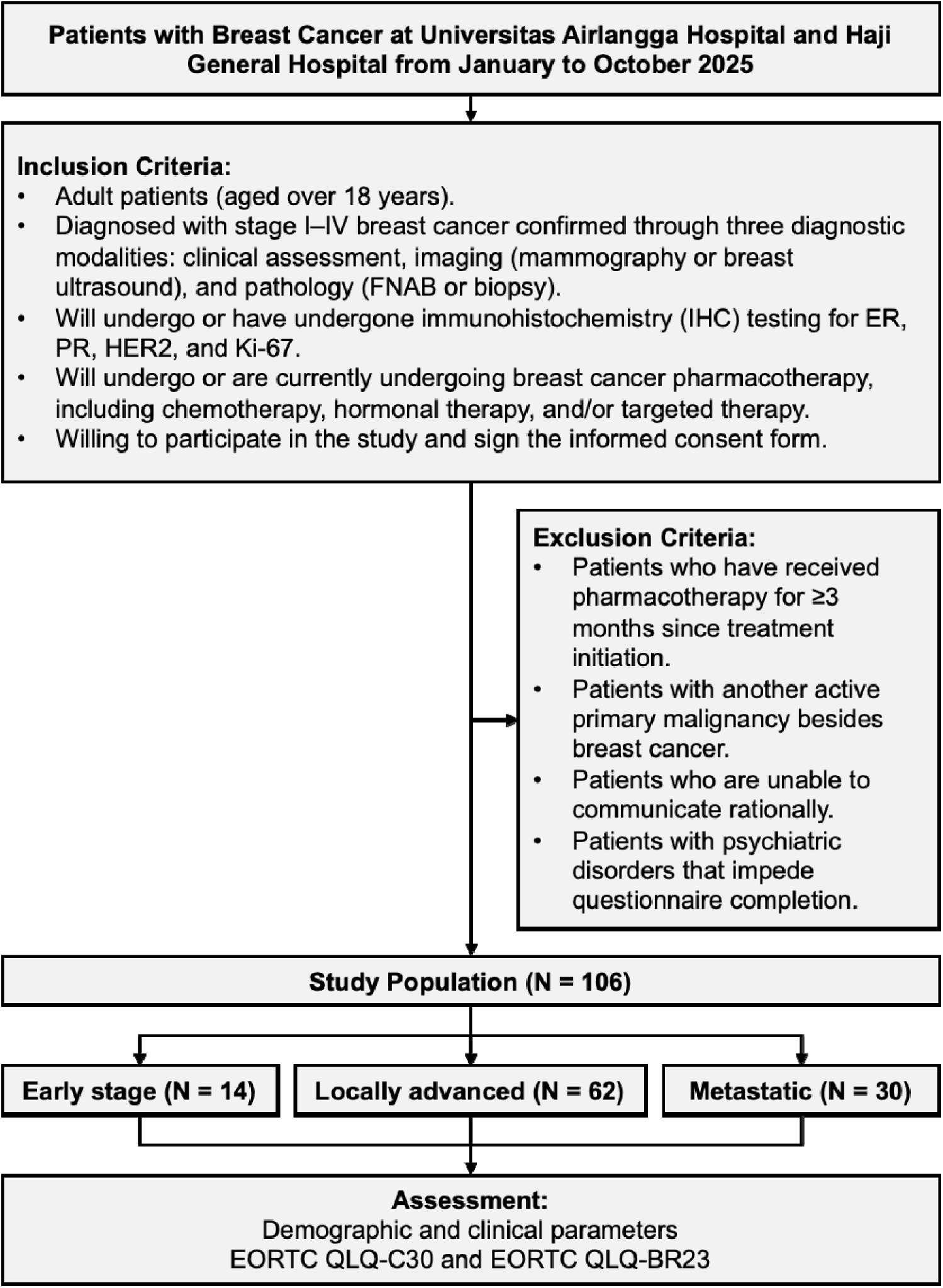
Flowchart of study design

### Clinical and histopathologic data collection

Baseline demographic information (age, sex, education level, employment status, marital status) and comorbidity profiles were collected during clinical interviews and chart review. Tumor characteristics—including histopathologic subtype, grade, and immunohistochemistry (IHC) profiles (ER, PR, HER2, and Ki67)—were extracted from pathology reports. Clinical staging was assigned using the American Joint Committee on Cancer (AJCC) 8^th^ edition TNM system. These data were used to classify patients into early-stage (stage I-IIA), locally advanced (stage IIB-III), or metastatic (stage IV) breast cancer groups, forming the basis of stage-stratified comparisons.

### QoL assessment

HRQoL was measured using the validated Indonesian-language versions of the EORTC QLQ-C30 and the breast cancer–specific module EORTC QLQ-BR23. The QLQ-C30 assesses global health status and functioning (physical, emotional, cognitive, role, social), as well as common cancer-related symptoms (fatigue, pain, nausea/vomiting, dyspnea, insomnia, appetite loss). The QLQ-BR23 captures breast cancer–specific concerns such as body image, sexual functioning, systemic therapy side effects, breast/arm symptoms, and distress related to hair loss. All scores were linearly transformed to a 0–100 scale according to EORTC scoring guidelines, with higher scores indicating better functioning or greater symptom burden, depending on the scale.

### Data management and quality control

Data were collected using standardized forms and entered into a secure, password-protected electronic database. Double-entry verification was performed by independent research assistants to minimize transcription errors. Missing questionnaire items were handled according to EORTC manual protocols, allowing score computation when ≥50% of items within a given domain were completed.

### Statistical analysis

Descriptive statistics were used to summarize baseline characteristics across disease stages. Continuous variables were presented as means with standard deviations or medians with interquartile ranges, depending on distribution assessed via Shapiro–Wilk testing. Categorical variables were summarized as frequencies and percentages. To examine differences in HRQoL across early, locally advanced, and metastatic breast cancer, the Kruskal–Wallis test was used for continuous, non-parametric domain scores. Statistical significance was set at *p* <0.05. All analyses were performed using SPSS version 25 (IBM Corp., Armonk, NY).

### Ethical considerations

The study complied with the Declaration of Helsinki and Good Clinical Practice guidelines. Ethical approval was granted by both institutional review boards prior to initiation. Participants received verbal and written explanations about the study purpose, procedures, and confidentiality safeguards, and informed consent was obtained before any data collection.

## Results

### Baseline characteristics of the study population

A total of 106 women with breast cancer were included in the analysis, comprising 14 patients with early-stage disease, 62 with locally advanced disease, and 30 with metastatic disease (**Table 1**). All participants across the three groups were female. The mean age differed slightly by stage, with early-stage patients being the oldest group (54.79 ± 9.1 years), followed by those with locally advanced disease (51.73 ± 9.9 years), and those with metastatic disease (50.97 ± 9.7 years). The distribution of educational attainment varied across the three groups. In the early-stage group, most patients had completed high school (42.9%) or higher education (28.6%). In contrast, the locally advanced group had a higher proportion of individuals with university-level education (45.2%), while elementary education accounted for 21.0%. The metastatic group showed a broader distribution, with 40.0% having completed high school, 23.3% having higher education, and 20.0% completing elementary school. Very few patients across all groups had no formal education. Employment patterns were similar across disease stages. In the early-stage group, 71.4% of patients were unemployed, while 28.6% were employed. Locally advanced disease demonstrated a slightly lower proportion of unemployment (61.3%), whereas the metastatic group had the highest proportion of unemployed participants (73.3%). Most patients across all stages were married. All patients with early-stage disease (100%) were married. Among the locally advanced group, 82.3% were married, 6.5% were single, and 11.3% were widowed. In the metastatic group, 86.7% were married and 13.3% were single, with no widowed patients recorded.

**Table 1.** Baseline characteristics of the study population.

| Parameter | Early<br>(N = 14) | Locally advanced<br>(N = 62) | Metastatic<br>(N = 30) |
| --- | --- | --- | --- |
| **Sex (n; %)** |  |  |  |
| Female | 14 (100.0%) | 62 (100.0%) | 30 (100.0%) |
| **Age in years (mean ± SD)** | 54.79 ± 9.1 | 51.73 ± 9.9 | 50.97 ± 9.7 |
| **Education (n; %)** |  |  |  |
| No formal education | 0 (0.0%) | 1 (1.6%) | 1 (3.3%) |
| Elementary | 3 (21.4%) | 13 (21.0%) | 6 (20.0%) |
| Secondary | 1 (7.1%) | 3 (4.8%) | 4 (13.3%) |
| High school | 6 (42.9%) | 17 (27.4%) | 12 (40.0%) |
| Higher education / university | 4 (28.6%) | 28 (45.2%) | 7 (23.3%) |
| **Employment status (n; %)** |  |  |  |
| Unemployed | 10 (71.4%) | 38 (61.3%) | 22 (73.3%) |
| Employed | 4 (28.6%) | 24 (38.7%) | 8 (26.7%) |
| **Marital status (n; %)** |  |  |  |
| Single | 0 (0.0%) | 4 (6.5%) | 4 (13.3%) |
| Married | 14 (100.0%) | 51 (82.3%) | 26 (86.7%) |
| Widow | 0 (0.0%) | 7 (11.3%) | 0 (0.0%) |
| **Comorbidities (n; %)** |  |  |  |
| **No** | 5 (35.7%) | 22 (42.9%) | 16 (53.3%) |
| **Yes:** | 9 (64.3%) | 40 (57.1%) | 14 (46.7%) |
| Hypertension (n) | 6 | 22 | 4 |
| Diabetes mellitus (n) | 0 | 5 | 7 |
| Cardiovascular diseases (n) | 2 | 8 | 6 |
| Respiratory diseases (n) | 3 | 3 | 1 |
| Miscellaneous (n) | 5 | 26 | 6 |
| **History of breast cancer treatment (n; %)** |  |  |  |
| **No** | 1 (7.1%) | 24 (38.7%) | 7 (23.3%) |
| **Yes** | 13 (92.9%) | 38 (61.3%) | 23 (76.7%) |
| Surgery (n) | 13 | 32 | 15 |
| Pharmacotherapy (n) | 6 | 20 | 23 |
| Radiotherapy (n) | 0 | 2 | 2 |

The prevalence of comorbidities differed among the groups. Early-stage patients had the highest proportion of comorbidities (64.3%), followed by the locally advanced group (57.1%) and the metastatic group (46.7%). Hypertension was the most frequently reported comorbidity in all stages, with six cases in early-stage, twenty-two in locally advanced, and four in metastatic disease. Diabetes mellitus was most prevalent among metastatic patients (n = 7), whereas cardiovascular disease was most common in the locally advanced group. Miscellaneous comorbidities—including conditions not classified under major organ systems—were reported in all groups, with the highest frequency in the locally advanced cohort. Most patients had a history of prior breast cancer-related treatment. Prior therapy was reported in 92.9% of early-stage, 61.3% of locally advanced, and 76.7% of metastatic patients. Surgical history was common, with nearly all early-stage patients (13 out of 14) having undergone surgery, compared with 32 patients in the locally advanced group and 15 in the metastatic group. Prior pharmacotherapy was reported in 6 early-stage, 20 locally advanced, and 23 metastatic patients. Radiotherapy was infrequently reported across all groups, noted in only two patients each in the locally advanced and metastatic categories, and none in the early-stage group.

### Breast cancer characteristics

The distribution of tumor histopathology varied across disease stages (**Table 2**). All patients with early-stage breast cancer had invasive ductal carcinoma (IDC) (100%), with no cases of invasive lobular carcinoma (ILC), mixed histology, or other subtypes recorded. In the locally advanced group, IDC remained the predominant histologic type (82.3%), followed by ILC (6.5%), mixed IDC–ILC tumors (6.5%), and a small number of miscellaneous subtypes (1.6%). Two cases (3.2%) had insufficient information and were categorized as unknown. Among patients with metastatic disease, IDC also constituted the majority (80.0%), while ILC and mixed tumors were observed in 3.3% and 13.3% of patients, respectively. One metastatic case (3.3%) was assigned to the miscellaneous category. Tumor grade distributions differed between early, locally advanced, and metastatic disease. Early-stage breast cancer demonstrated a balanced distribution between grade II (42.9%) and grade III tumors (42.9%), with grade I tumors accounting for 7.1% of cases and one case (7.1%) classified as unknown. In the locally advanced cohort, grade III tumors were the most common (48.4%), followed by grade II (30.6%) and grade I (9.7%); seven cases (11.3%) had no available grading information. In patients with metastatic disease, grade II tumors were most frequent (43.3%), followed by grade III (23.3%) and grade I (10.0%). The metastatic cohort also had seven cases (23.3%) categorized as unknown grade, representing the highest proportion of missing grading data among the three groups.

**Table 2.** Breast cancer profile of the study population.

| Parameter | Early<br>(N = 14) | Locally advanced<br>(N = 62) | Metastatic<br>(N = 30) |
| --- | --- | --- | --- |
| **Tissue histopathology (n; %)** |  |  |  |
| Invasive ductal carcinoma (IDC) | 14 (100.0%) | 51 (82.3%) | 24 (80.0%) |
| Invasive lobular carcinoma (ILC) | 0 (0.0%) | 4 (6.5%) | 1 (3.3%) |
| Mixed type of IDC and ILC | 0 (0.0%) | 4 (6.5%) | 4 (13.3%) |
| Miscellaneous (e.g., *mucinous*) | 0 (0.0%) | 1 (1.6%) | 0 (0.0%) |
| Unknown | 0 (0.0%) | 2 (3.2%) | 1 (3.3%) |
| **Tumor grade (n; %)** |  |  |  |
| Grade I | 1 (7.1%) | 6 (9.7%) | 3 (10.0%) |
| Grade II | 6 (42.9%) | 19 (30.6%) | 13 (43.3%) |
| Grade III | 6 (42.9%) | 30 (48.4%) | 7 (23.3%) |
| Unknown | 1 (7.1%) | 7 (11.3%) | 7 (23.3%) |
| **Immunohistochemistry (n; %)** |  |  |  |
| Luminal A | 3 (21.4%) | 4 (6.5%) | 3 (10.0%) |
| Luminal B HER2- | 3 (21.4%) | 17 (27.4%) | 8 (26.7%) |
| Luminal B HER2+ | 2 (14.3%) | 11 (17.7%) | 7 (23.3%) |
| HER2 *enriched* | 4 (28.6%) | 11 (17.7%) | 1 (3.3%) |
| HER2 *low* | 1 (7.1%) | 3 (4.8%) | 3 (10.0%) |
| TNBC | 1 (7.1%) | 5 (8.1%) | 2 (6.7%) |
| Unknown | 0 (0.0%) | 11 (17.7%) | 6 (20.0%) |

IHC subtypes demonstrated variability across disease stages. Among early-stage patients, Luminal A and Luminal B (HER2–) each accounted for 21.4% of cases. Luminal B (HER2+) and HER2-enriched subtypes constituted 14.3% and 28.6% of cases, respectively. HER2-low and triple-negative breast cancer (TNBC) each represented 7.1% of the early-stage group, with no unknown cases reported. In the locally advanced group, Luminal B (HER2–) was the most common subtype (27.4%), followed by Luminal B (HER2+) and HER2-enriched subtypes, each accounting for 17.7%. Luminal A made up a smaller proportion (6.5%). HER2-low tumors were identified in 4.8% of patients, and TNBC in 8.1%. Eleven patients (17.7%) had incomplete IHC results and were classified as unknown. In the metastatic group, Luminal B (HER2–) remained the most frequent subtype (26.7%), followed by Luminal B (HER2+) (23.3%). Luminal A and HER2-low subtypes each accounted for 10.0% of metastatic cases, whereas HER2-enriched tumors were observed in 3.3% of patients. TNBC was identified in 6.7% of metastatic patients. The metastatic cohort also had a notable proportion of cases (20.0%) with unknown IHC data.

### QoL comparison across disease stages

Global health status scores showed variation across disease stages (**Table 3**). Patients with early-stage breast cancer reported the highest median score of 83.3 (IQR 81.3–93.8), followed by those with locally advanced disease with a median of 83.3 (IQR 66.7–91.7). Metastatic patients had the lowest median global health status score of 75.0 (IQR 47.9–83.3). The difference among the three groups was statistically significant (*p* = 0.004). Physical function scores were highest in early-stage disease (median 100.0, IQR 91.7–100.0) and locally advanced disease (median 100.0, IQR 80.0–100.0), while metastatic patients demonstrated lower scores (median 86.7, IQR 63.3–100.0). Although progressively lower scores were observed with advancing stage, the difference did not reach statistical significance (*p* = 0.068). Role function scores remained high across all stages. Early-stage and locally advanced groups had identical median scores of 100.0, with narrow IQRs. Metastatic disease showed a wider distribution (median 100.0, IQR 66.7–100.0). The observed differences were not statistically significant (*p* = 0.067). Emotional function scores were similar across stages, with all groups having a median score of 83.3. Interquartile ranges were widest in metastatic disease (50.0–100.0), but overall differences were not significant (*p* = 0.696). Cognitive function scores were comparable across early, locally advanced, and metastatic disease groups, each showing a median of 100.0 and overlapping IQRs. No significant difference was observed (*p* = 0.990). Social functioning showed greater variability. Early-stage patients had a median score of 100.0 (IQR 100.0–100.0), similar to locally advanced disease (median 100.0, IQR 95.8–100.0). Metastatic patients had lower scores (median 100.0, IQR 62.5–100.0). The difference reached statistical significance (*p* = 0.022).

**Table 3.** Comparison of quality of life domains across stages.

| Parameter | Early<br>Median [IQR]<br>(N = 14) | Locally advanced<br>Median [IQR]<br>(N = 62) | Metastatic<br>Median [IQR]<br>(N = 30) | p-value<sup>#</sup> |
| --- | --- | --- | --- | --- |
| **Global health status** |  |  |  |  |
| Quality of life (QL2) | 83.3 [81.3 – 93.8] | 83.3 [66.7 – 91.7] | 75.0 [47.9 – 83.3] | **0.004** |
| **C30 – Functional scale** |  |  |  |  |
| Physical function (PF2) | 100.0 [91.7 – 100.0] | 100.0 [80.0 – 100.0] | 86.7 [63.3 – 100.0] | 0.068 |
| Role function (RF2) | 100.0 [100.0 – 100.0] | 100.0 [95.8 – 100.0] | 100.0 [66.7 – 100.0] | 0.067 |
| Emotional function (EF) | 83.3 [66.7 – 100.0] | 83.3 [66.7 – 91.7] | 83.3 [50.0 – 100.0] | 0.696 |
| Cognitive function (CF) | 100.0 [83.3 – 100.0] | 100.0 [83.3 – 100.0] | 100.0 [83.3 – 100.0] | 0.990 |
| Social function (SF) | 100.0 [100.0 – 100.0] | 100.0 [95.8 – 100.0] | 100.0 [62.5 – 100.0] | **0.022** |
| **C30 – Symptom scale** |  |  |  |  |
| Fatigue (FA) | 22.2 [0.0 – 33.3] | 22.2 [0.0 – 44.4] | 33.3 [8.3 – 58.3] | 0.178 |
| Nausea vomiting (NV) | 0.0 [0.0 – 16.7] | 0.0 [0.0 – 0.0] | 0.0 [0.0 – 0.0] | 0.597 |
| Pain (PA) | 16.7 [0.0 – 33.3] | 16.7 [12.5 – 50.0] | 33.3 [16.7 – 50.0] | **0.041** |
| Dyspnea (DY) | 0.0 [0.0 – 8.3] | 0.0 [0.0 – 0.0] | 0.0 [0.0 – 33.3] | 0.704 |
| Insomnia (SL) | 33.3 [0.0 – 66.7] | 0.0 [0.0 – 66.7] | 33.3 [0.0 – 66.7] | 0.878 |
| Appetite loss (AP) | 0.0 [0.0 – 0.0] | 0.0 [0.0 – 33.3] | 0.0 [0.0 – 33.3] | 0.098 |
| Constipation (CO) | 0.0 [0.0 – 0.0] | 0.0 [0.0 – 0.0] | 0.0 [0.0 – 33.3] | 0.332 |
| Diarrhea (DI) | 0.0 [0.0 – 0.0] | 0.0 [0.0 – 0.0] | 0.0 [0.0 – 0.0] | 0.196 |
| Financial issue (FI) | 0.0 [0.0 – 0.0] | 0.0 [0.0 – 33.3] | 0.0 [0.0 – 41.7] | 0.150 |
| **BR23 – Functional scale** |  |  |  |  |
| Body image (BRBI) | 100.0 [81.3 – 100.0] | 91.7 [83.3 – 100.0] | 95.8 [66.7 – 100.0] | 0.571 |
| Sexual function (BRSEF) | 33.3 [0.0 – 33.3] | 33.3 [16.7 – 33.3] | 16.7 [0.0 – 33.3] | 0.062 |
| Sexual enjoyment (BRSEE) | 16.7 [0.0 – 33.3] | 33.3 [0.0 – 33.3] | 33.3 [0.0 – 33.3] | 0.087 |
| Future perspective (BRFU) | 50.0 [25.0 – 100.0] | 66.7 [33.3 – 100.0] | 66.7 [33.3 – 100.0] | 0.972 |
| **BR23 – Symptom scale** |  |  |  |  |
| Systemic side effects (BRST) | 4.8 [0.0 – 10.7] | 4.8 [3.6 – 19.0] | 14.3 [8.3 – 23.8] | **0.025** |
| Breast symptom (BRBS) | 12.5 [8.3 – 18.8] | 8.3 [0.0 – 33.3] | 16.7 [8.3 – 25.0] | 0.867 |
| Arm symptom (BRAS) | 11.1 [0.0 – 33.3] | 11.1 [0.0 – 33.3] | 22.2 [8.3 – 44.4] | 0.238 |
| Upset by hair loss (BRHL) | 0.0 [0.0 – 0.0] | 0.0 [0.0 – 0.0] | 0.0 [0.0 – 0.0] | 0.976 |

<sup>#</sup> Comparison of three independent groups using Kruskal-Wallis independent sample test

Fatigue scores increased progressively with disease stage: early (median 22.2), locally advanced (22.2), and metastatic (33.3). However, the difference did not reach statistical significance (*p* = 0.178). Nausea and vomiting scores were low across all stages, with median scores of 0.0 in all groups. The early-stage group displayed a slightly wider IQR (0.0–16.7). No statistically significant difference was observed (*p* = 0.597). Pain scores increased with advancing stage. Early-stage patients reported a median of 16.7 (IQR 0.0–33.3), similar to the locally advanced group (16.7, IQR 12.5–50.0). Metastatic patients showed higher scores (33.3, IQR 16.7–50.0). The difference between groups was statistically significant (*p* = 0.041). Dyspnea scores were low in all groups, with medians of 0.0. The metastatic group had a wider IQR (0.0–33.3), yet differences remained non-significant (*p* = 0.704). Insomnia scores demonstrated variability but no significant difference across stages. Medians were 33.3 in early and metastatic disease and 0.0 in locally advanced disease (*p* = 0.878). Appetite loss scores were generally low, with median values of 0.0 in all groups. Differences were not statistically significant (*p* = 0.098). Constipation scores were uniformly low across all stages, with median 0.0 and no significant difference (*p* = 0.332). Diarrhea scores were also low, with median 0.0 in all groups and no significant variation (*p* = 0.196). Financial difficulty scores were low across stages with no significant differences (*p* = 0.150). IQRs were wider in metastatic patients, indicating greater variability.

Body image scores were highest in early-stage disease (median 100.0), slightly lower in metastatic (95.8), and lowest in locally advanced disease (91.7). Differences were not statistically significant (*p* = 0.571). Sexual function scores showed decreasing medians across stages—33.3 in early and locally advanced disease and 16.7 in metastatic—but without significant differences (*p* = 0.062). Sexual enjoyment scores were also similar, with medians of 16.7, 33.3, and 33.3 across the three groups (*p* = 0.087). Future perspective scores were relatively similar across all stages, with medians ranging from 50.0 to 66.7. Differences were not significant (*p* = 0.972). Systemic therapy-related symptom scores were lowest in early-stage disease (median 4.8) and highest in metastatic disease (median 14.3). Locally advanced disease had intermediate scores (4.8). The difference among groups reached statistical significance (*p* = 0.025). Breast symptoms and arm symptoms were comparable across the three groups, with no significant differences (*p* = 0.867 and *p* = 0.238, respectively). Medians ranged between 8.3–16.7 for breast symptoms and 11.1–22.2 for arm symptoms. Scores for distress related to hair loss were uniformly 0.0 across all stages, with no observed variation (*p* = 0.976).

## Discussion

The baseline characteristics of this cohort demonstrate several important patterns that reflect both global breast cancer epidemiology and region-specific realities of patient presentation. All participants were female, which is expected given the overwhelmingly higher incidence of breast cancer in women [9]. The mean age across all stages—ranging from approximately 51 to 55 years—is consistent with the observation that breast cancer in many Asian and LMIC populations tends to occur at a younger age compared with Western cohorts, where peak incidence typically arises between 60 and 70 years [10]. The slightly younger mean age in locally advanced and metastatic groups may suggest delays in detection and diagnosis among middle-aged women, a trend commonly attributed to low screening participation, socioeconomic barriers, and the absence of population-level mammography programs in many developing regions [11].

Educational attainment varied across stages, with a higher proportion of university-educated patients in the locally advanced group and a more heterogeneous distribution in metastatic disease. These findings highlight the complex interplay between health literacy, socioeconomic status, and healthcare-seeking behaviors [12]. While higher education is generally associated with earlier detection and better outcomes, the predominance of locally advanced rather than early-stage disease even among those with higher education suggests that structural barriers—such as limited screening access, cultural taboos regarding breast health, or diagnostic delays—may overshadow individual-level knowledge. Conversely, the more diverse educational background in metastatic patients may reflect cumulative disparities in health awareness and healthcare navigation that contribute to late presentation.

Employment status across the cohort shows a predominance of unemployment, particularly among early-stage and metastatic patients. Unemployment in breast cancer cohorts is often multifactorial: patients may already experience functional limitations or psychological distress at diagnosis, or they may be unemployed due to socioeconomic realities predating cancer. The slightly higher unemployment rate in metastatic disease is expected, as advanced cancer is frequently accompanied by symptomatic burden, mobility restrictions, and financial strain that can hinder employment [13]. The observation that early-stage patients also demonstrate high unemployment may reflect broader socioeconomic patterns within the population rather than disease-specific effects. Marital status showed the expected predominance of married individuals across all stages, reflecting demographic norms in many Asian countries [14]. However, the presence of widowed or single individuals predominantly in the locally advanced group may have indirect implications. Lack of spousal support has been associated with delays in symptom recognition, reduced screening engagement, and higher psychological burden [15]. These social determinants may contribute to more advanced disease at presentation, although causality cannot be inferred.

The prevalence of comorbidities in this cohort—particularly hypertension, diabetes mellitus, and cardiovascular disease—is consistent with the rising burden of non-communicable diseases in the region. Comorbid conditions were most frequent in early-stage disease and least frequent in metastatic disease, a pattern potentially explained by survivor bias and selective recruitment. Patients with multiple comorbidities may have more frequent healthcare encounters, facilitating earlier detection of breast symptoms. Conversely, advanced-stage patients may represent a subset of individuals with lower engagement in healthcare systems, fewer routine screenings, or socioeconomic obstacles limiting early detection despite comorbidity presence [16]. The metabolic and inflammatory milieu associated with hypertension and diabetes has also been proposed to influence tumor progression and treatment tolerance, suggesting that comorbidity profiles may have prognostic implications in later analyses [17]. Prior treatment history differed substantially between stages. Nearly all early-stage patients had undergone prior surgery, consistent with standard curative-intent management. In locally advanced and metastatic groups, the lower proportion of prior surgery and higher use of pharmacotherapy reflect both clinical staging and treatment algorithms: advanced disease is typically managed with neoadjuvant systemic therapy or palliative pharmacotherapy rather than upfront surgical intervention. The higher proportion of metastatic patients with prior pharmacotherapy is expected, as systemic therapy is the backbone of treatment in metastatic disease. The sparse use of radiotherapy across all stages likely reflects treatment sequencing at the time of data collection, variable access to radiation facilities, or timing relative to the baseline QoL assessment.

The distribution of tumor histopathology in this cohort aligns with global epidemiologic patterns, with IDC comprising the vast majority of cases across all stages [18]. IDC accounted for 100% of early-stage tumors and remained the predominant subtype in locally advanced (82.3%) and metastatic disease (80.0%). This dominance is consistent with international data reporting IDC as the most common breast cancer histology, representing approximately 70–80% of cases worldwide. The slightly higher frequency of mixed IDC–ILC tumors in metastatic disease (13.3%) may reflect the inherently infiltrative, discohesive biology of lobular components, which are known to disseminate more diffusely and may be underdiagnosed at initial presentation [19]. Additionally, the presence of miscellaneous histologies, though limited, highlights the heterogeneity of breast cancer in real-world settings and underscores the importance of detailed pathological classification in guiding therapy. Tumor grade patterns across disease stages provide important insights into disease biology. Grade II and III tumors dominated all stages, with high-grade tumors being proportionally greatest in locally advanced disease (48.4%). This observation is consistent with the well-established association between higher grade and more aggressive tumor behavior, including faster proliferation, increased genomic instability, and reduced differentiation [20]. The high proportion of grade III tumors in the locally advanced group may suggest more rapid tumor progression or delayed detection, both of which could contribute to patients presenting with larger tumor burden or nodal involvement. Meanwhile, metastatic patients had a relatively higher proportion of grade II tumors, a pattern sometimes seen in real-world cohorts where the histologic grade may not fully predict metastatic behavior, particularly in tumors driven by specific molecular pathways or receptor phenotypes. The substantial proportion of unknown grades in metastatic patients (23.3%) also reflects real-world limitations in tissue availability, sampling constraints, or incomplete diagnostic workups, particularly when metastasis is diagnosed clinically or radiographically.

The IHC subtype distribution reflects the molecular diversity of breast cancer and has important implications for prognosis and therapy. Luminal B (HER2– and HER2+) subtypes were the most common across locally advanced and metastatic disease, consistent with the global trend of Luminal B tumors presenting at more advanced stages due to their higher proliferative index (e.g., Ki-67) and more aggressive clinical course compared with Luminal A tumors [21]. The relatively modest proportion of Luminal A tumors in locally advanced (6.5%) and metastatic (10.0%) groups is expected, as Luminal A cancers typically exhibit slower growth and are more likely to be detected at earlier stages, often through screening. Their greater presence in early-stage disease (21.4%) parallels international observations that screen-detected tumors are frequently Luminal A. HER2-enriched tumors were most common in early-stage disease (28.6%) and less frequent in the metastatic setting (3.3%). This pattern may reflect improved early detection of symptomatic HER2-positive lesions due to their rapid growth, or may indicate successful earlier-stage treatment reducing progression into the metastatic category. Conversely, HER2-low tumors—recently recognized as a distinct biologic group with therapeutic relevance—were found across all stages but remained a minority subtype [22]. The emergence of HER2-low classification in this cohort underscores the growing importance of precise HER2 quantification, particularly in the era of antibody–drug conjugates. The distribution of triple-negative breast cancer (TNBC) was relatively low in all groups, though slightly higher in locally advanced (8.1%) and metastatic disease (6.7%) compared with early-stage (7.1%). While TNBC is often associated with advanced-stage presentation due to its rapid proliferation and lack of targeted therapy options, the modest representation of TNBC in this cohort may reflect population-specific differences in molecular epidemiology [23]. However, it also emphasizes that non-TNBC aggressive subtypes, such as Luminal B and HER2-enriched tumors, substantially contribute to advanced-stage burden in this population. The proportion of unknown IHC subtypes—17.7% in locally advanced and 20.0% in metastatic disease—likely reflects practical issues such as limited access to tissue sampling, incomplete receptor testing, or referral patterns where diagnostic workups are performed at multiple institutions. These gaps highlight the continuing challenge of ensuring uniform biomarker evaluation in resource-limited settings, which remains essential for appropriate therapeutic planning.

The stage-stratified assessment of QoL in this cohort reveals several important patterns that reflect both the biological progression of breast cancer and the psychosocial challenges associated with advanced disease [24]. Global health status declined significantly with increasing stage, with metastatic patients reporting the lowest scores. This finding mirrors global evidence demonstrating that tumor burden, symptom intensity, and functional impairments typically accumulate as cancer progresses. The wide interquartile range among metastatic patients further highlights the heterogeneity in how individuals experience late-stage disease—some maintaining stable functioning while others experience profound decline. Together, these results affirm that disease stage remains a strong determinant of overall perceived health even before treatment begins. Functional domain analyses provide additional clarity on how advancing disease impacts daily functioning. Physical function showed a downward trend from early to metastatic disease, consistent with the increasing presence of pain, fatigue, systemic inflammation, and organ dysfunction commonly associated with advanced tumors. Although role function remained high overall, its wider variability in metastatic patients suggests that while some individuals maintain their capacity to perform daily roles, others experience substantial disruption. Emotional and cognitive function remained relatively stable across stages, a pattern also seen in several international QoL studies, suggesting that psychological resilience and coping strategies may remain intact despite clinical progression—though this interpretation warrants caution given the cross-sectional design. Notably, social functioning showed a significant decline with advancing stage, reflecting the impact of physical symptoms, mobility limitations, hospital visits, and emotional withdrawal on patients’ ability to participate in social activities.

Symptom burden demonstrated clear stage-dependent patterns. Pain scores increased significantly with advancing disease, which is expected given the higher prevalence of tumor infiltration, skeletal metastasis, neuropathic involvement, and inflammation in metastatic cancer [25]. This finding aligns with global reports identifying pain as one of the most prevalent and debilitating symptoms in metastatic breast cancer [26]. Fatigue, while not statistically different across stages, showed a clear upward trend, consistent with disease-related metabolic changes, anemia, systemic inflammation, and psychological distress that commonly intensify with advanced disease. Nausea, vomiting, dyspnea, constipation, and diarrhea were uniformly low across all stages, which is expected given that the assessments were conducted prior to systemic therapy, when many treatment-induced toxicities have not yet emerged. Financial difficulties, although not significantly different between groups, showed greater variability in metastatic patients. This reflects the broader socioeconomic impact of advanced cancer, including loss of income, increased healthcare visits, and ancillary costs of supportive care. Similar trends have been widely documented in global cancer survivorship literature, particularly in LMIC settings where out-of-pocket expenses may contribute to financial toxicity.

Breast cancer–specific QoL domains from the QLQ-BR23 further illustrate how advancing stage affects patient well-being. Body image scores were high across all groups, though slightly lower in patients with locally advanced disease—potentially reflecting psychological responses to visible breast changes, ulceration, or preoperative tumor bulk. Sexual function and sexual enjoyment were generally low, with metastatic patients reporting the lowest scores. These findings align with existing evidence showing that sexual health is often compromised by physical symptoms, emotional strain, and body image disturbances, particularly in advanced disease [27,28]. Systemic therapy–related side effects, while assessed before treatment initiation, were significantly higher in metastatic patients. These symptoms may reflect disease-related systemic manifestations, including fatigue, appetite changes, and generalized discomfort, as well as residual toxicities from past treatments in patients with relapsed or recurrent disease. Breast and arm symptoms were comparable across stages, suggesting that localized symptoms may be driven more by tumor location or prior surgery than by stage progression. Distress associated with hair loss was uniformly absent across all stages, consistent with the pre-treatment timing of the assessment.

### Clinical Insights and Future Direction

The findings from this study provide important clinical insights into how breast cancer stage at presentation shapes the patient experience even before systemic therapy begins. While prior studies have documented stage-dependent differences in outcomes such as survival or treatment response, fewer have comprehensively described the intersection between sociodemographic background, tumor biology, and multidimensional patient-reported QoL in a real-world LMIC referral setting. This dataset reveals a consistent pattern: advanced disease correlates with younger age at presentation, more aggressive tumor biology (particularly Luminal B subtypes and high-grade carcinomas), and significantly worse global health status, social functioning, pain, and systemic symptom burden. These findings underscore that the challenges faced by patients with advanced breast cancer begin long before the first dose of therapy, emphasizing the need for earlier detection, multidisciplinary support, and proactive symptom management strategies tailored to stage-specific vulnerabilities.

What is particularly novel in this cohort is the integration of detailed biological profiling with robust HRQoL assessment, allowing a more comprehensive understanding of how tumor characteristics translate into lived patient experience. The combination of high unemployment, varied educational attainment, and substantial comorbidity burden suggests that social determinants of health may amplify the clinical impact of disease stage. Meanwhile, the high representation of aggressive molecular subtypes in advanced disease highlights the dual burden of delayed detection and unfavorable tumor biology. These insights suggest that interventions must extend beyond improving access to treatment; they must also address the structural and psychosocial barriers that shape when and how patients seek care. Incorporating early supportive care, psychosocial counseling, and pain management—beginning at diagnosis rather than after treatment initiation—may meaningfully improve patient trajectories. Moving forward, this evidence supports several actionable directions. First, strengthening early detection pathways—including community-based education, risk communication, and low-cost clinical breast examinations—may help shift stage distribution toward earlier diagnosis [29]. Second, integrating routine HRQoL screening into baseline oncology evaluations can enable clinicians to identify high-risk individuals who would benefit from early palliative involvement or social support interventions [30]. Third, addressing gaps in biomarker testing and pathology completeness, particularly in advanced disease, could reduce therapeutic delays and allow more precise treatment selection [31]. Finally, future research should examine how baseline QoL interacts with treatment tolerance, hematologic toxicity, and survival outcomes in this population, helping refine personalized care pathways. By bridging biological, clinical, and psychosocial dimensions, these findings provide a foundation for developing comprehensive, stage-tailored care models that improve both QoL and clinical outcomes in breast cancer patients across resource settings.

## Conclusion

In this cohort of women with breast cancer, advancing disease stage was associated with more aggressive tumor biology and marked deterioration in key domains of health-related QoL even before systemic therapy was initiated. Patients with locally advanced and metastatic disease experienced significantly lower global health status, reduced social functioning, and higher pain and systemic symptom burdens, reflecting the combined impact of tumor progression, sociodemographic factors, and unmet supportive care needs. These findings highlight the urgency of improving early detection pathways and underscore the value of integrating routine quality-of-life assessment and stage-tailored supportive interventions into baseline oncology care. Strengthening these approaches may help mitigate symptom burden, enhance treatment readiness, and optimize patient-centered outcomes across the breast cancer continuum.

## Data Availability

All data produced in the present study are available upon reasonable request to the authors

## Notes

### Competing Interest Statement

The authors have declared no competing interest.

### Funding Statement

This study did not receive any funding

### Author Declarations

Ethics committee of Universitas Airlangga Hospital and Haji General Hospital gave ethical approval for this work.

